# Potent induction of humoral and cellular immunity after bivalent BA.4/5 mRNA vaccination in dialysis patients with and without history of SARS-CoV-2 infection

**DOI:** 10.1101/2023.08.13.23294045

**Authors:** Saskia Bronder, Janine Mihm, Rebecca Urschel, Verena Klemis, Tina Schmidt, Stefanie Marx, Amina Abu-Omar, Franziska Hielscher, Candida Guckelmus, Marek Widera, Urban Sester, Martina Sester

**Author notes:** Correspondence: Martina Sester, PhD, Saarland University, Department of Transplant and Infection Immunology, Institutes of Infection Medicine, building 47, Kirrberger Straße, D-66421 Homburg, Germany.

## Abstract

Knowledge on immunogenicity of the bivalent Omicron BA.4/5 vaccine in dialysis patients and the effect of a previous infection is limited. Therefore, vaccine-induced humoral and cellular immunity was analyzed in dialysis patients and immunocompetent controls with and without prior infection.

In an observational study, 33 dialysis patients and 58 controls matched for age, sex and prior infection status were recruited. Specific IgG, neutralizing antibody activity and cellular immunity towards the spike-antigen from parental SARS-CoV-2 and Omicron subvariants BA.1, BA.2 and BA.4/5 were analyzed before and 13-18 days after vaccination.

The bivalent vaccine led to a significant induction of IgG, neutralizing titers, and specific CD4 and CD8 T-cell levels. Neutralizing activity towards the parental strain was highest, whereas specific T-cell levels towards parental spike and Omicron subvariants did not differ indicating substantial cross-reactivity. Dialysis patients with prior infection had significantly higher spike-specific CD4 T-cell levels with lower CTLA-4 expression compared to infection-naïve patients. When compared to controls, no differences were observed between individuals without prior infection. Among infected individuals, CD4 T-cell levels were higher in dialysis patients and neutralizing antibodies were higher in controls. Vaccination was overall well tolerated in both dialysis patients and controls with significantly less adverse events among dialysis patients.

In conclusion, our study did not provide any evidence for impaired immunogenicity of the bivalent Omicron BA.4/5 vaccine in dialysis patients. Unlike in controls, previous infection of patients was even associated with higher levels of spike-specific CD4 T cells, which may reflect prolonged encounter with antigen during infection.

**Translational statement:** Dialysis patients with uremic immunodeficiency are at increased risk for infectious complications after SARS-CoV-2 infection and have been shown to insufficiently respond towards the first doses of COVID-19 vaccines. Bivalent vaccines are now recommended, although knowledge on immunogenicity and on the effect of a previous infection is limited in this patient group. We show that the bivalent BA.4/5 vaccine was well tolerated and led to a pronounced induction of antibodies, neutralizing antibodies and T cells, which was overall similar in magnitude in non-infected patients and controls. Despite some differences between patients and controls with prior infection, our data do not provide any evidence towards impaired immunity in dialysis patients.

## Introduction

End-stage chronic kidney disease (CKD) and dialysis are risk factors for severe acute respiratory syndrome coronavirus 2 (SARS-CoV-2) infection and the development of severe COVID-19 disease with fatal outcome which was particularly evident at the beginning of the pandemic ^1^. This resulted from the relative inability to perform stringent physical distancing due to regular hemodialysis treatment and associated travel to and from the dialysis centers ^2^ as well as from general immunological impairments due to uremic immunodeficiency, advanced age, and multiple comorbidities ^3^. To protect this vulnerable group from severe COVID-19 disease, hospitalizations, and death, patients were prioritized for COVID-19 vaccination once licensed. In line with a decreased response rate to vaccinations such as hepatitis B, tetanus ^3, 4^, or influenza ^5^, patients were shown to frequently mount an inadequate humoral and cellular immune response after COVID-19 vaccination ^6–10^ which waned rapidly after administration of prime and booster doses ^11, 12^. This necessitated more frequent booster vaccinations in this vulnerable patient group to achieve a similar level of immunity as in immunocompetent individuals. In 2022, the immune-escaping omicron subvariants of concern dominated the pandemic, which led to an increased incidence of breakthrough infections in both patients and immunocompetent individuals. This is illustrated by the fact that four doses of the monovalent mRNA vaccine resulted in a high neutralizing activity against the ancestral strain, but a much lower rate against the omicron subvariant BA.1 ^13^, BA.4 and BA.5 ^14^. Therefore, bivalent mRNA vaccines targeting both the parental strain and either BA.1 or BA.4/5 were developed to more specifically induce variant-adapted humoral and cellular immunity. First studies among immunocompetent controls showed a more pronounced induction of neutralizing antibodies after bivalent booster vaccination compared to the monovalent vaccination ^15–18^. The bivalent BA.4/5 vaccine effectiveness was 72% for preventing COVID-19 hospitalization, and 68% for preventing COVID-19-related deaths compared to individuals who did not receive a booster vaccination ^19^. With increasing incidence of breakthrough infection, knowledge on the effect of hybrid immunity on subsequent booster vaccinations becomes increasingly important. We and others have previously shown that the induction of humoral immunity after bivalent vaccination in immunocompetent individuals is more pronounced in previously non-infected individuals ^15, 16, 20^, whereas vaccine-induced T-cell levels were similar in both groups ^20^. Similar data on bivalent vaccines in dialysis patients are limited. First immunogenicity data are available that were either restricted to the analysis of humoral immunity ^21^ or did not differentiate between individuals with and without prior infection ^22^. Moreover, no head-to-head analyses with immunocompetent controls are available.

We therefore prospectively characterized induction of humoral and cellular immunity after bivalent BA.4/5 vaccination in dialysis patients. Patients with and without prior infection were compared regarding spike-specific IgG, neutralizing titers as well as CD4 and CD8 T-cell levels against parental SARS-CoV-2 and variants of concern. Moreover, immunogenicity and reactogenicity of the bivalent BA.4/5 booster vaccination were compared between patients and age-matched immunocompetent controls.

## Methods

### Study design and subjects

In this observational study, continuous ambulatory peritoneal dialysis and hemodialysis patients were enrolled prior to their vaccination with the bivalent BA.4/5 vaccine (Comirnaty® Original/Omicron BA.4/5, BioNTech/Pfizer). A subgroup of healthy immunocompetent (HC) volunteers (mainly employees at Saarland University Medical campus), were included as controls. Study participants received a questionnaire for self-reporting their history of vaccination and infection, and of local and systemic adverse events within the first week after vaccination. Heparinized blood samples were collected before and 13-18 days after vaccination to determine specific humoral and cellular immunity toward the spike protein derived from the parental SARS-CoV-2 strain as well as from the Omicron variants BA.1, BA.2 and BA.4/5. The study was approved by ethics committee of the Ärztekammer des Saarlandes (reference 76/20 including amendment), and written consent was obtained from all individuals.

### Quantification of vaccine-induced spike-specific T cells

To determine spike-specific T cells, heparinized whole blood was stimulated for 6h as described before ^23, 24^ with overlapping peptides (each peptide 2 µg/ml) spanning the parental spike or Omicron variants BA.1-, BA.2-, BA.4/5-spike protein (N-terminal receptor binding domain and C-terminal portion including the transmembrane domain, jpt Berlin, Germany) in the presence of co-stimulatory antibodies against CD28 and CD49d (clone L293 and clone 9F10, 1 μg/ml each). In addition, stimulation with 0.64% DMSO and 2.5 μg/ml of *Staphylococcus aureus* enterotoxin B (SEB; Sigma) served as a negative and positive control, respectively. After stimulation, cells were immunostained using anti-CD4 (clone SK3, 1:33.3), anti-CD8 (clone SK1, 1:12.5), anti-CD69 (clone L78, 1:33.3), anti-IFNγ (clone 4S.B3, 1:100), anti-IL-2 (clone MQ1-17H12, 1:12.5), anti-TNFα (clone MAb11, 1:20), and anti-CTLA-4 (clone BNI3, 1:50) and analyzed using flow cytometry (BD FACS Canto II and FACSDiva software 6.1.3.). Activated CD69-positive T cells producing IFNγ identified spike (WT, BA.1, BA.2, BA.4/5)-reactive CD4 or CD8 T cells. Levels of reactive CD4 and CD8 T cells after control stimulations were subtracted from those obtained after spike-specific stimulation, and 0.03% of reactive T cells was set as detection limit as described before ^23^. To characterize T-cell functionality, co-expression of IL-2 and TNFα was analyzed as well as the cytotoxic T-lymphocyte-associated Protein 4 (CTLA-4).

### UV-inactivated viral strains

In this study, the following SARS-CoV-2 isolates were used: Parental strain (SARS-CoV-2 B.1 FFM7/2020, GenBank ID MT358643), BA.1 (SARS-CoV-2 B.1.1.529 FFM-SIM0550/2021 (EPI_ISL_6959871), GenBank ID OL800702), BA.2 (SARS-CoV-2 BA.2 FFM-BA.2-3833/2022, GenBank ID OM617939), BA.5 (SARS-CoV-2 BA.5 FFM-BA.5-501/2022, GenBank ID OP062267) ^25–29^. For use in neutralization assays, the strains were UV-inactivated as described previously ^30, 31^.

### Determination of SARS-CoV-2-specific antibodies

All antibody tests were performed according to the manufacturer’s instructions (Euroimmun, Lübeck, Germany) as described before ^23^. The enzyme-linked immunosorbent assay (ELISA, SARS-CoV-2-QuantiVac) was used to quantify the SARS-CoV-2-specific IgG antibodies towards the receptor binding domain of the parental SARS-CoV-2-spike protein. Antibody binding units (BAU/ml) <25.6 were scored negative, ≥25.6 and <35.2 were scored intermediate, and ≥35.2 were scored positive. SARS-CoV-2-specific IgG towards the nucleocapsid (NCP) protein were determined using the anti-SARS-CoV-2-NCP-ELISA (Euroimmun, Lübeck, Germany). NCP-ELISA positivity was used as independent evidence for infection in individuals without history of infection. A micro neutralization assay with A549-AT cells and authentic parental SARS-CoV-2 (FFM7, D614G) and the Omicron variants BA.1, BA.2, and BA.5 was used to determine the in vitro neutralizing activity of the antibodies, as described before ^28, 31^.

### Statistical analysis

All statistical analyses were performed using GraphPad Prism 10.0.2.232 software (GraphPad, San Diego. CA, USA) using two-tailed tests. Categorial analyses on sex and adverse events were performed using Fisher’s exact test. Data with normal distribution were analyzed using unpaired t test. To compare unpaired nonparametric data between groups, Mann-Whitney and Kruskal–Wallis test followed by Dunn’s multiple comparisons test were performed. Wilcoxon matched pairs test was used to compare paired data between two groups. Correlations were analyzed using a correlation matrix according to Spearman. A p-value less than 0.05 was considered statistically significant.

## Results

### Study population

Thirty-three patients undergoing hemodialysis (n=32) or continuous ambulatory peritoneal dialysis (n=1) were recruited, of which 17 had a history of a prior SARS-CoV-2 infection. In addition, despite no reported history of SARS-CoV-2 infection, two additional patients were assigned to the infection group due to a positive nucleocapsid-specific IgG. All patients were tested prior to and at a median of 16 (IQR 2) days after vaccination with the bivalent BA.4/5 vaccine. In both groups, most patients had a history of homologous mRNA vaccination with at least two and up to five prior immunization events (including vaccinations and infections, figure 1A). The patients with prior infection were younger and had a longer time on dialysis than the infection-naïve patients (table 1). Demographic characteristics including primary disease that led to renal failure resulting in dialysis treatment, comorbidities, and differential blood counts are shown in table 1. The number of leukocytes and granulocytes did not differ between the groups, whereas infection-naïve patients showed a significantly higher number of monocytes (p=0.011) and lymphocytes (p=0.023, table 1).

**Figure 1.**
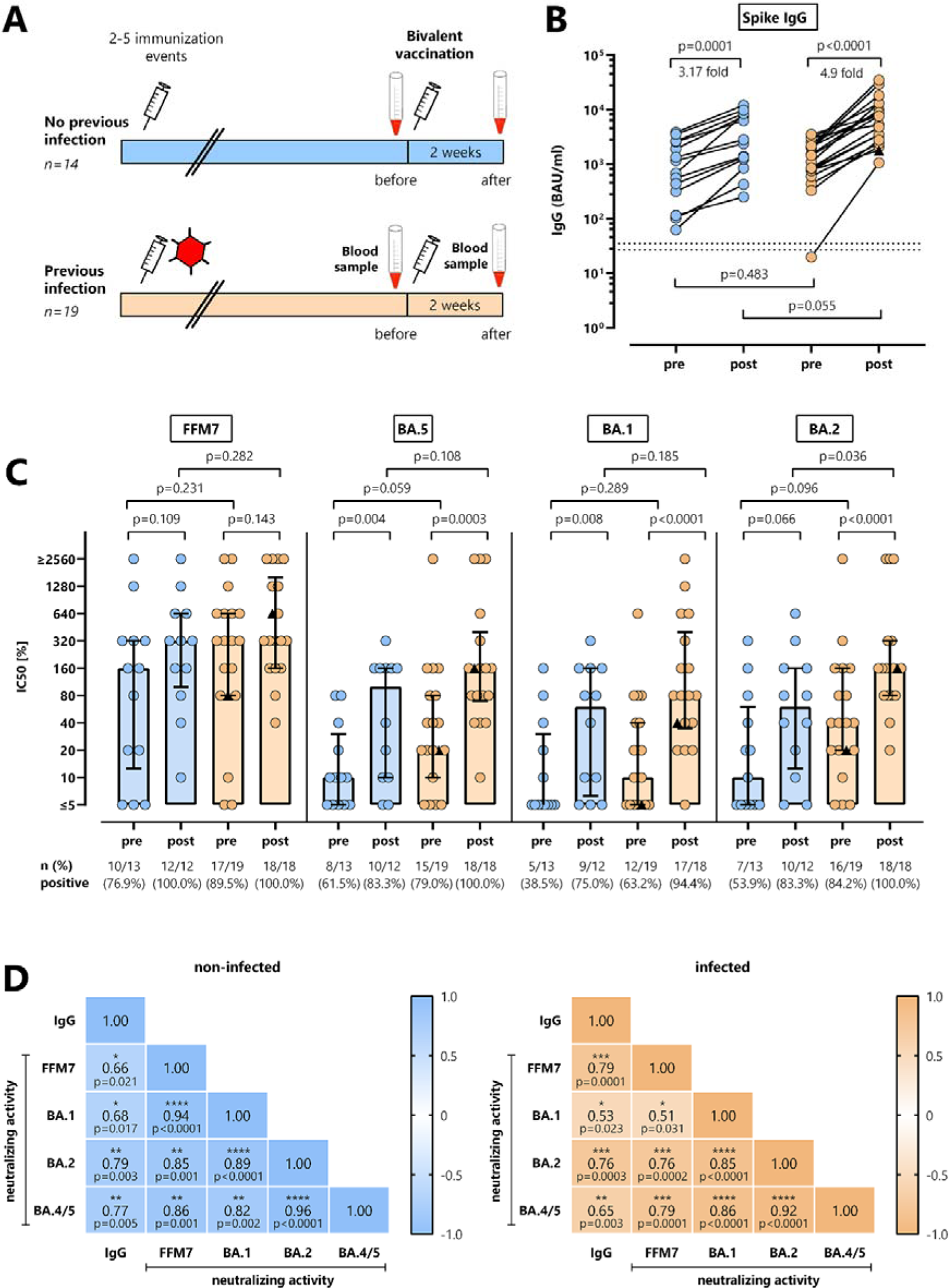
Bivalent BA.4/5 vaccine-induced antibodies levels and neutralizing activity in dialysis patients with and without prior infection. **(A)** Schematic outline of the study design, where blood was drawn dialysis patients without (n=14, blue) and with prior infection (n=19; orange) before and after bivalent BA.4/5 vaccination. **(B)** Levels of spike-specific IgG antibodies towards parental spike protein (expressed as BAU/ml) were analyzed at baseline and after vaccination. Lines represent medians with interquartile ranges. **(C)** Antibody-mediated neutralization of parental SARS-CoV-2 (FFM7) and Omicron BA.1, BA.2 and BA.5 variants of concern (expressed as IC _50_) was tested among dialysis patients with and without prior infection as well as before and after vaccination with the bivalent BA.4/5 vaccine. Bars represent medians with interquartile ranges. Differences were calculated using Wilcoxon signed rank test (before/after) or Mann-Whitney test for group comparisons at baseline and after vaccination. **(D)** Correlation between IgG levels towards parental spike and neutralizing activity towards parental SARS-CoV-2 (FFM7) and Omicron BA.1, BA.2 and BA.5 variants of concern are displayed in a correlation matrix. Correlations coefficients were calculated according to two-tailed Spearman and displayed using a color code. One patient, marked by a black triangle, was infected with SARS-CoV-2 two days after bivalent vaccination.

**Table 1.** Demographic and clinical characteristics of the study population of dialysis patients.

|  | non-infected<br>n=14 | infected<br>n=19 | p-value |
| --- | --- | --- | --- |
| Years of age, mean (SD) | 71.8 (11.3) | 58.7 (15.5) | 0.012<sup>§</sup> |
| Sex, n (%)<sup>†</sup> |  |  |  |
| female | 7 (50.0%) | 5 (26.3%) | 0.273<sup>†</sup> |
| male | 7 (50.0%) | 14 (73.7%) |  |
| Vaccine regimen, n (%) |  |  |  |
| mRNA | 13 (92.9%) | 17 (89.5%) |  |
| Vector/mRNA combination | 1 (7.1%) | 2 (10.5%) |  |
| Total number of prior immunization events, n (%) |  |  |  |
| 2 | 1 (7.1%) | 1 (5.3%) |  |
| 3 | 8 (57.1%) | 10 (52.6%) |  |
| 4 | 5 (35.7%) | 6 (31.6%) |  |
| 5 | 0 (0%) | 2 (10.5%) |  |
| Type of latest immunization prior to vaccination with Comirnaty BA.4/5, n (%) |  |  |  |
| infection | n.a. | 4 (21.1%) |  |
| vaccination | 14 (100.0%) | 13 (68.4%) |  |
| unknown |  | 2 (10.5%)* |  |
| Analysis time<br>(days after vaccination), median (IQR) | 16 (2) | 16 (2) |  |
| Type of dialysis, n (%) |  |  |  |
| peritoneal dialysis | 1 (7.1%) | - |  |
| hemodialysis | 13 (92.9%) | 19 (100.0%) |  |
| Time on dialysis<br>(years), mean (SD) | 1.48 (1.61) | 5.25 (4.09) | 0.003<sup>§</sup> |
| Cause of kidney failure, n (%) |  |  |  |
| autoimmune-mediated<br>nephropathy | 2 (15.4%) | 1 (5.3%) |  |
| chronic glomerulonephritis | 1 (7.1%) | 3 (15.8%) |  |
| secondary chronic renal<br>disease | 10 (71.4%) | 9 (47.4%) |  |
| innate | 1 (7.1%) | 5 (26.3%) |  |
| unknown |  | 1 (5.3%) |  |
| Comorbidities (n) |  |  |  |
| Diabetes mellitus | 7 | 10 |  |

|  | non-infected | infected | p-value |
| --- | --- | --- | --- |
|  | n=14 | n=19 |  |
| diabetes mellitus | 3 | 5 |  |
| arterial hypertension | 11 | 18 |  |
| coronary heart disease | 4 | 6 |  |
| lung disease |  | 3 |  |
| liver cirrhosis |  | 2 |  |
| active tumor | 1 | 1 |  |
| ANCA-associated<br>vasculitides |  | 1 |  |
| Differential blood counts | n=13 | n=19 |  |
| median (IQR) cells/μl | | | |
| Leukocytes | 6600 (4550) | 5800 (1900) | 0.420<sup>‡</sup> |
| Granulocytes | 4891 (2536) | 4437 (1764) | 0.821<sup>‡</sup> |
| Monocytes | 704 (266) | 536 (130) | 0.011<sup>‡</sup> |
| Lymphocytes | 1280 (663) | 1051 (480) | 0.023<sup>‡</sup> |

<sup>§</sup>unpaired t-test, <sup>†</sup>Fisher's exact test, <sup>‡</sup>Mann-Whitney test, <sup>†</sup>information on sex was based on individual self-declaration; \*2 patients were assigned as infected based on NCAP-IgG positivity despite no known history of infection. IQR, interquartile range

### SARS-CoV-2-specific antibodies after bivalent vaccination in patients with and without prior infection

Blood samples were drawn prior to and a median of 16 (IQR 2) days after vaccination (figure 1A). IgG towards the parental spike were detectable in 32/33 patients prior to vaccination with no difference between individuals with and without prior infection (p=0.483). Both groups showed a significant induction of IgG after vaccination with no difference in the relative increases (figure 1B, p=0.0001, 3.17-fold and p<0.0001, 4.9-fold increase in non-infected and infected individuals, respectively). Despite similar baseline IgG-levels, vaccine-induced median IgG-levels in patients with prior infection were slightly higher (7689 (IQR 10217) BAU/ml) than in patients without prior infection (2604 (IQR 7011) BAU/ml), although the difference did not reach statistical significance (p=0.055, figure 1B). Antibodies were further characterized for their neutralizing ability towards the spike proteins targeted by the vaccine (parental strain (FFM7) and omicron BA.5) as well as Omicron BA.1 and BA.2 (figure 1C). Baseline neutralizing titers against the parental strain were considerably high in both infected and non-infected patients, and only slightly increased after vaccination. In contrast, baseline neutralizing titers against the Omicron variants BA.5, BA.1 and BA.2 were lower, and significantly increased in both non-infected and infected patient groups. Both the percentage of individuals with detectable neutralizing antibodies as well as median titers reached after vaccination were slightly higher in infected than in non-infected patients. Overall, IgG levels and neutralizing antibody activity towards the parental strain and all omicron subvariants showed a significant correlation in both patient groups (figure 1D).

### SARS-CoV-2-specific CD4 and CD8 T-cell levels after bivalent vaccination in patients with and without prior infection

Characterization of the spike-specific cellular immune response before and after vaccination was performed after stimulation with overlapping peptide pools derived from the parental spike protein followed by intracellular cytokine staining. In addition, SEB-stimulation was used to analyze polyclonal T-cell responses. Spike-specific T cells were identified by co-expression of CD69 and IFNγ, and both vaccine-induced CD4 and CD8 T-cell levels towards parental spike exceeded the detection limit in the majority of cases (figure 2A). The vaccine induced a significant increase in spike-specific CD4 T-cell levels in both non-infected (1.76-fold, p=0.014) and infected patients (1.86-fold, p=0.006) Likewise, median percentages of spike-specific CD8 T cells showed a significant increase (2.90-fold in infection-naïve (p=0.004) and 2.64-fold in infected patients (p=0.008)). Except from a significant increase in SEB-reactive CD4 T cells among non-infected patients, polyclonal CD4 and CD8 T-cell levels did show any pronounced vaccine-induced dynamics (figure 2B).

**Figure 2.**
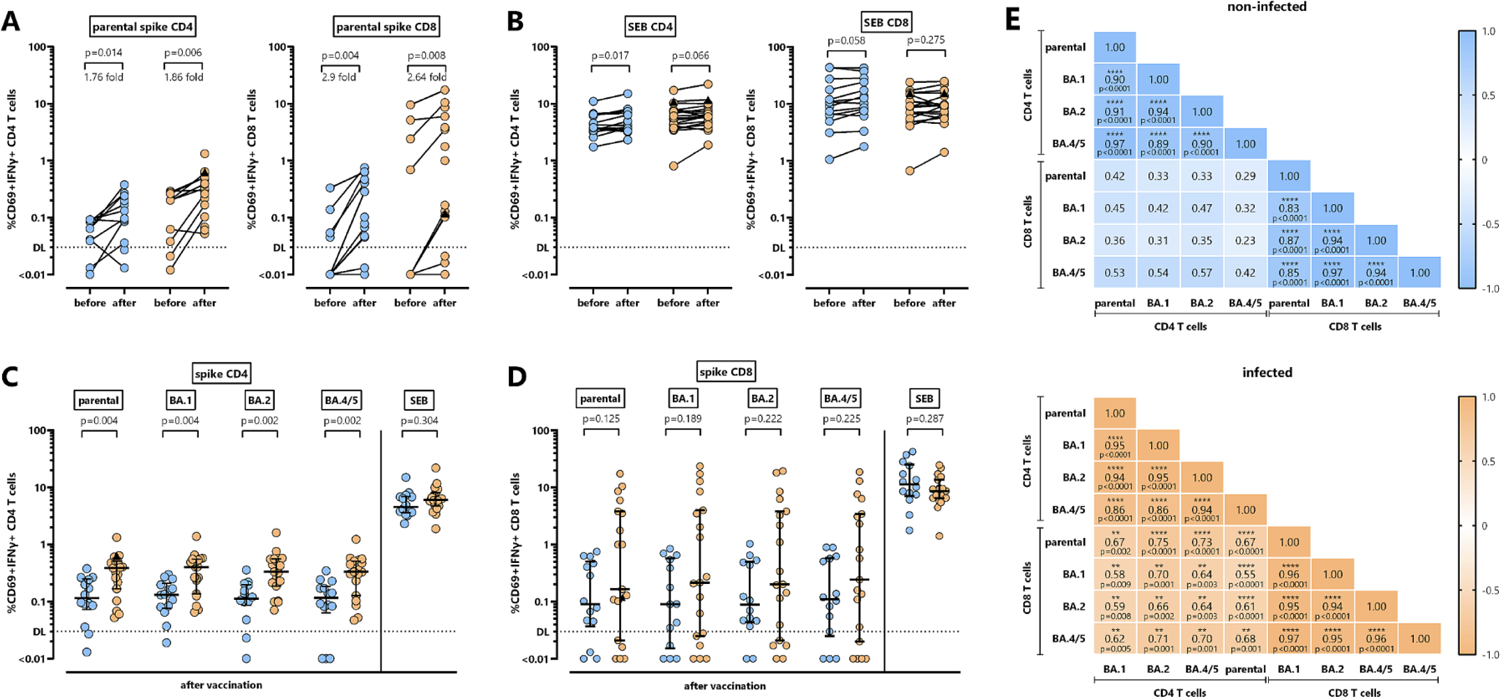
Bivalent BA.4/5 vaccine-induced CD4 and CD8 T-cell immunity in dialysis patients with and without prior infection. **(A)** CD4 and CD8 T cells towards parental spike were determined at baseline (n=10 in patients without and with prior infection) and after vaccination in dialysis patients without (n=14, blue symbols) and with prior infection (n=19; orange symbols). Fold changes after vaccination are indicated above the graphs and were calculated by dividing the individual levels after vaccination and levels prior to vaccination (with 0.03% added to each value prior to division to avoid division by 0). **(B)** SEB-reactive CD4 and CD8 T cells were determined at baseline and after vaccination in dialysis patients without (n=14) and with prior infection (n=19). **(C)** CD4 and **(D)** CD8 T cells towards parental spike and towards spike from Omicron subvariants BA.1, BA.2, BA.4/5, and SEB-reactive CD4 and CD8 T cells were compared after vaccination of patients without (n=14) and with prior infection (n=19). One patient, marked by a black triangle, was infected with SARS-CoV-2 two days after vaccination. Bars represents medians with interquartile ranges. Differences between time points were calculated by Wilcoxon signed rank test (A) and between the groups using Mann-Whitney test (B, C). Dotted lines indicate detection limits (DL) for spike-specific CD4 and CD8 T cells. **(E)** Correlation matrix of CD4 and CD8 T-cell levels towards parental spike and Omicron subvariants BA.1, BA.2 and BA.4/5 (n=33 dialysis patients). Correlation coefficients were calculated according to two-tailed Spearman and displayed using a color code.

When comparing the specific CD4 and CD8 T cells towards spike from the parental strain and Omicron subvariants between non-infected and infected patients after vaccination, the median percentage of parental spike-specific CD4 T cells was lower in infection-naïve patients (figure 2C), whereas spike-specific CD8 T-cell levels did not differ between the two groups (figure 2D). Likewise, there was no significant difference in median percentages of SEB-reactive CD4 and CD8 T cells between the two groups (figure 2C and D). Interestingly, the bivalent vaccine also induced specific CD4 and CD8 T cells towards all Omicron subvariants, which not only included Omicron BA.4/5 as part of the vaccine, but also Omicron BA.1 and BA.2. As with specific CD4 T cells towards parental spike, BA.1, BA.2 and BA.4/5-specific CD4 T-cell levels were significantly higher among infected than in patients without infection (figure 2C). In addition, among both CD4 and CD8 T cells, there was a significant correlation between spike-specific T-cell levels towards the parental spike and all tested Omicron subvariants BA.1, BA.2 and BA.4/5 (figure 2E). Interestingly, as shown in the lower left part of the correlation matrices, significant correlations between specific CD4 and CD8 T-cell populations were only found among individuals with prior infection (figure 2E).

### Functional and phenotypical characteristics of bivalent BA.4/5 vaccine-induced T cells of patients with and without prior infection

Apart from quantitative analyses of spike-specific CD4 and CD8 T cells after vaccination, which was based on the induction of IFNγ, the phenotypical and functional characteristics of specific T cells were further evaluated by cytokine profiling. Individual or combined expression of IFNγ, TNFα and IL-2 was characterized by Boolean gating, allowing the distinction of seven subpopulations which included polyfunctional cells simultaneously expressing all three cytokines, two cytokines, or one cytokine only. As shown in figure 3A and B, the cytokine profile of spike-specific CD4 and CD8 T cells was clearly distinct from SEB-reactive T cells. The majority of spike-specific CD4 T cells were polyfunctional, followed by dual-positive cells expressing TNFα in combination with either IFNγ or IL-2. The percentage of triple positive polyfunctional CD4 T cells towards parental spike was slightly higher among patients with prior infection, whereas non-infected patients had slightly higher levels of TNFα ^+^ IL-2 ^+^ dual positive CD4 T cells towards spike from BA.4/5 (figure 3A). Spike-specific CD8 T cells produced less IL-2 and were predominantly IFNγ ^+^ TNFα ^+^. In addition, a similar cytokine profile was observed for specific CD4 or CD8 T cells towards the Omicron subvariants BA.1 and BA.2 (supplementary figure S1).

**Figure 3.**
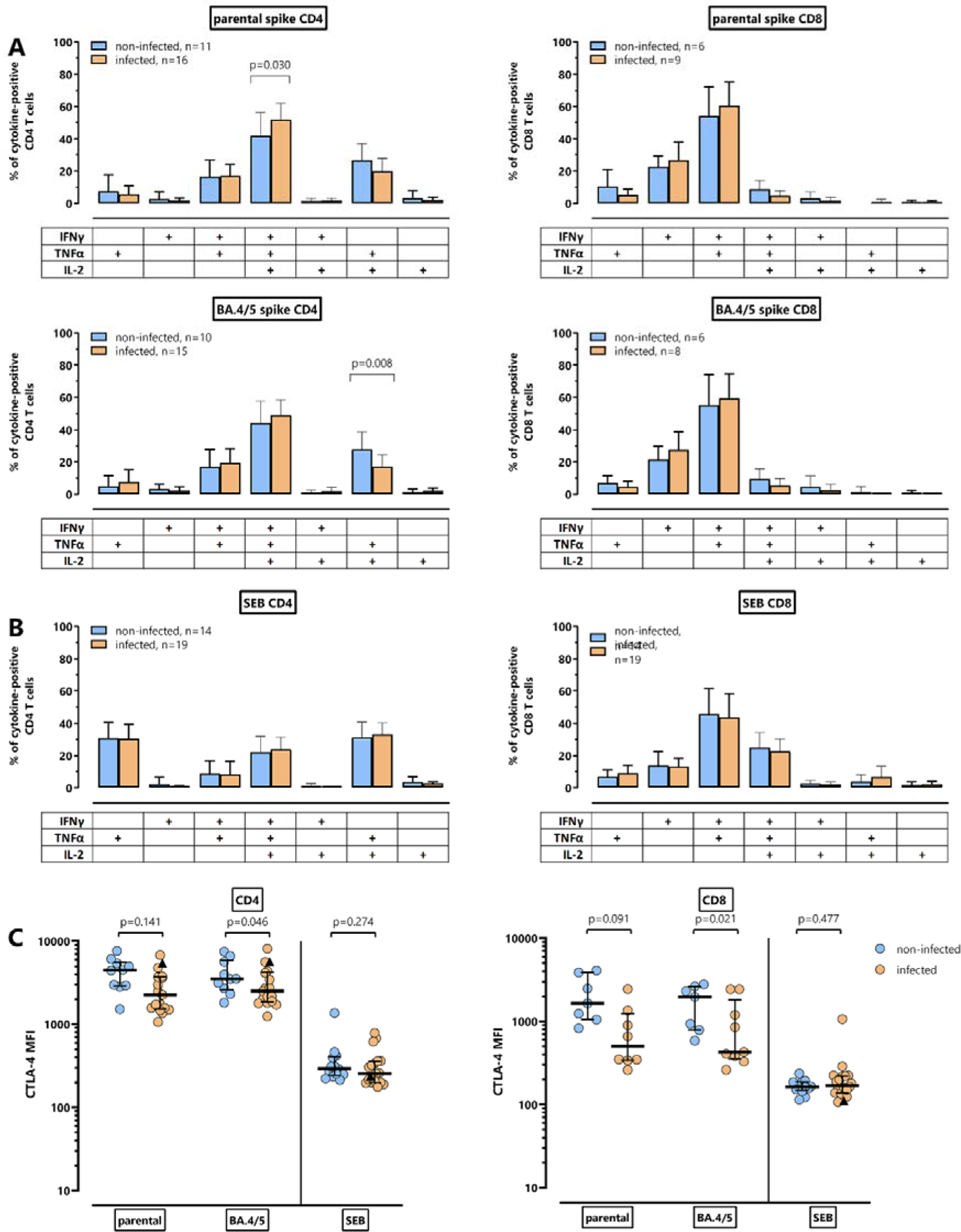
Functional characterization of parental and Omicron BA.4/5-spike-specific CD4 and CD8 T cells after bivalent vaccination. Cytokine expression profiles of CD4 and CD8 T cells after stimulation with **(A)**parental and BA.4/5-spike-peptides or **(B)** *Staphylococcus aureus* enterotoxin B (SEB) were compared between patients without (blue bars) and with (orange bars) prior infection. At the single-cell level, the cytokine-expressing T cells were differentiated into 7 subpopulations according to their expression of IFN-γ, TNF-α and IL-2 (single, double or triple cytokine-expressing cells). Only samples of the patients with at least 30 cytokine-expressing CD4 or CD8 T cells were included, respectively to allow for robust statistical analysis. Bars represent means and standard deviations of subpopulations. Differences among subpopulations were determined using unpaired t-test. **(C)** Median fluorescence intensity (MFI) of CTLA-4 expression on CD4 and CD8 T cells towards spike from the parental strain or BA.4/5, and on SEB-reactive CD4 and CD8 T cells from patients with and without prior infection was determined. To allow robust statistical analysis, only samples with at least 20 cytokine-positive CD4 or CD8 T cells, respectively, were included. Differences between groups were analyzed using Mann-Whitney test.

As evidence for recent encounter with antigen, spike-specific and SEB-reactive CD4 and CD8 T cells from infected and non-infected patients were compared regarding their CTLA-4 expression (figure 3C). CTLA-4 expression levels on spike-specific CD4 and CD8 T cells in infection-naïve patients were numerically higher than in patients with prior infection with statistically significant differences observed for spike-specific CD4 and CD8 T cells towards Omicron BA.4/5. These differences in CTLA-4 expression were spike-specific, as CTLA-4 expression on SEB-reactive CD4 and CD8 T cells were similarly low in both groups.

### Comparison of vaccine-induced humoral and cellular immunity in hemodialysis patients and immunocompetent controls

To evaluate potential differences in vaccine-induced immunity in patients and individuals without immunodeficiency, patient data were compared with those of 58 immunocompetent individuals who were matched for age, sex and prior infection status. Demographic characteristics and differential blood counts are shown in supplementary table S1. While the number of granulocytes and monocytes did not differ between patients and controls, leukocyte counts among individuals with prior infection were significantly lower in patients than in controls. Moreover, irrespective of prior infection, lymphocyte counts were significantly lower in patients than in controls.

Irrespective of prior infection, spike-specific IgG levels after vaccination were induced to a similar extent in both patients and controls (figure 4A). The neutralizing activity among individuals without prior infection was also similar in patients and controls. Interestingly, among individuals with prior infection, median neutralizing titers were numerically higher in immunocompetent controls, with significant differences for titers towards the parental strain and towards BA.2 (figure 4B). Comparison of vaccine-induced parental and Omicron subvariant-specific T cells showed no significant difference in spike-specific CD4 T cells between infection-naïve patients and controls (figure 4C). Interestingly, however, while spike-specific CD4 T-cell levels among controls was similar irrespective of prior infection, dialysis patients with prior infection showed a significantly higher percentage of spike-specific CD4 T cells than controls (figure 4C). This was different from CD8 T cells, where there was no difference between patients and controls, which held true for both individuals with and without prior infection (figure 4D). Although SEB-reactive CD4 T-cell responses among infected individuals were also slightly higher in patients than in controls (p=0.001), polyclonal T-cell responses were generally higher and rather of similar magnitude in all groups (figure 4C and D).

**Figure 4.**
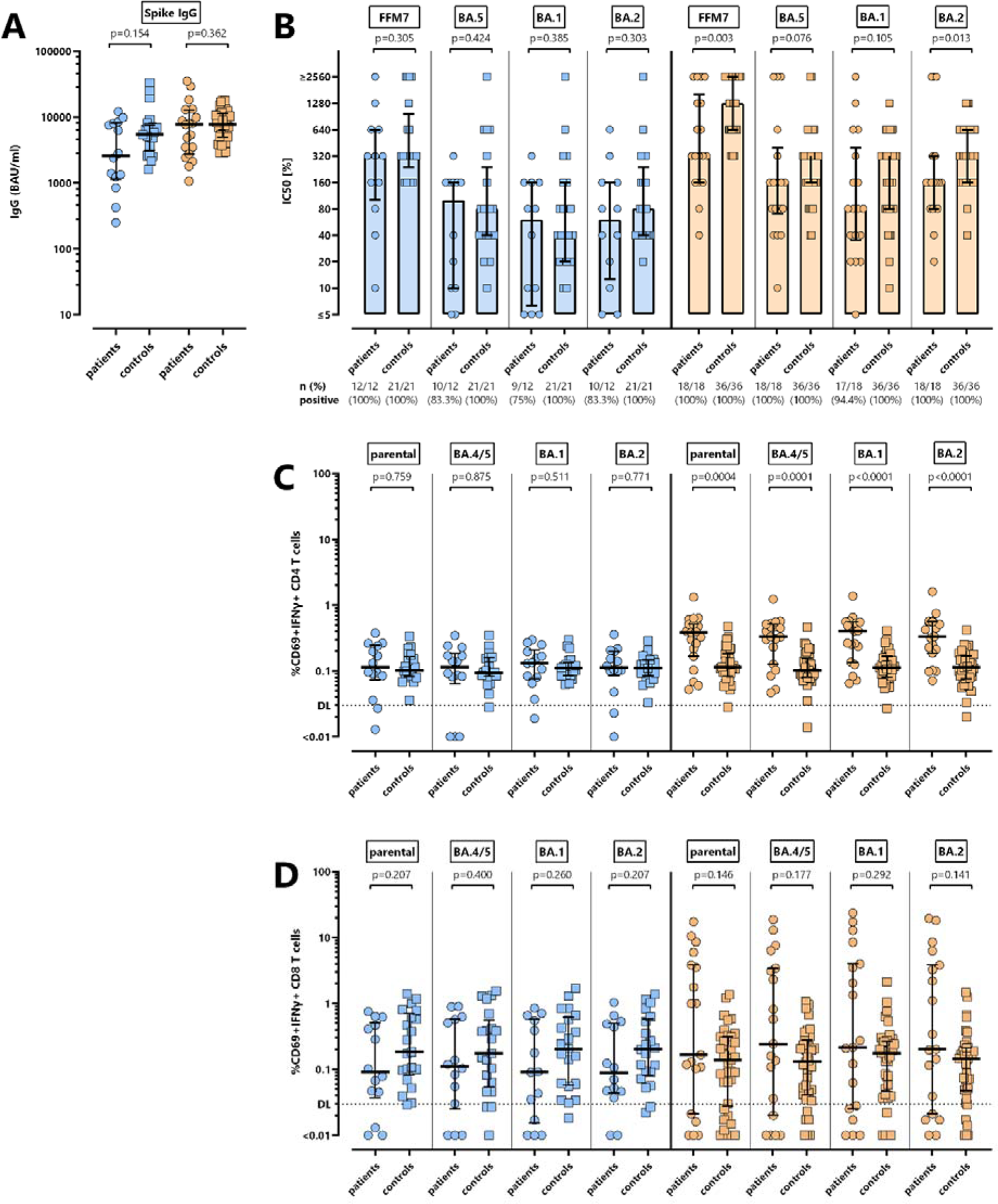
Comparison of bivalent BA.4/5 vaccine-induced humoral and cellular immunity between dialysis patients and healthy controls stratified according to their infection status. **(A)** Spike-specific IgG levels after bivalent vaccination were compared between infection-naïve dialysis patients (n=14, blue circle) and controls (n=21, blue square) as well as between previously infected dialysis patients (n=19, orange dots) and controls (n=37; orange squares). **(B)** Antibody-mediated neutralization of parental SARS-CoV-2 strain (FFM7) and Omicron BA.1, BA.2 and BA.5 variants of concern after vaccination with the bivalent vaccine (expressed as IC _50_) was compared between dialysis patients and controls with and without prior infection. The number of tested individuals and the percentage of individuals with detectable neutralizing antibodies are indicated. Levels of **(C)** CD4 and **(D)** CD8 T cells towards spike from the parental strain and Omicron subvariants BA.4/5, BA.1, BA.2 after vaccination were compared in patients and controls with and without prior infection. Dotted lines indicate detection limits (DL) for spike-specific CD4 and CD8 T cells. Bars represent median with interquartile ranges. Differences between the groups were determined using Mann-Whitney test.

### Reactogenicity after bivalent vaccination in patients and controls

Finally, self-reported local and systemic adverse events within the first week after vaccination were compared between patients and controls using a questionnaire. Irrespective of prior infection status, the vaccine was well tolerated among dialysis patients with most reporting either no (>50%) or only local adverse events at the injection site (figure 5A). Systemic adverse events included chills and gastrointestinal symptoms, but were only rarely reported among patients. In contrast, irrespective of prior infection status, local (mainly pain at the injection site) and systemic adverse events (mainly fatigue) were significantly more frequently reported among controls (figure 5A and B). In light of these rather minor adverse events of the bivalent vaccination, it is notable that all previous COVID-19 vaccinations were also very well tolerated in patients, as adverse events were all perceived as of similarly low severity. This was significantly different from controls, where a larger fraction felt most affected by the bivalent vaccination (figure 5C).

**Figure 5.**
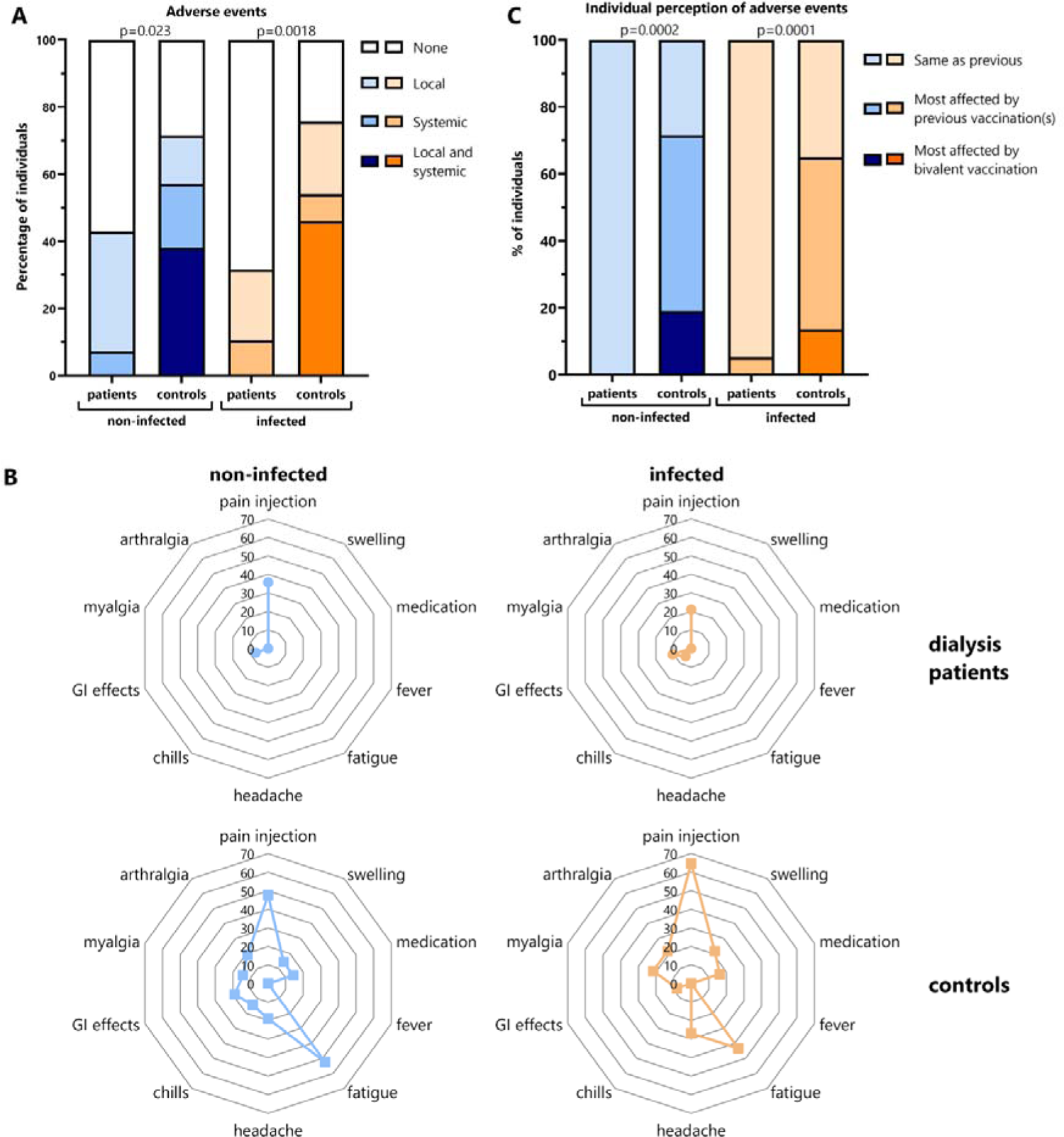
Reactogenicity after bivalent BA.4/5 vaccination in dialysis patients and controls. Reactogenicity within first week after bivalent BA.4/5 vaccination of dialysis patients and controls with and without prior infection was self-reported based on a standardized questionnaire. **(A)** The percentage of individuals with no adverse events, only local or only systemic adverse events or both are shown. **(B)** The distribution of the individual local and systemic adverse events in infected and non-infected patients and controls is shown. **(C)** Individual perception of adverse events were classified as to whether individuals felt most affected by the bivalent BA.4/5 vaccination, by any of the previous vaccinations, or whether adverse events were similar for all vaccinations. Comparisons between groups were analyzed using the X^2^ test.

## Discussion

Bivalent COVID-19 vaccines have been recommended for use as booster doses in both immunocompetent and immunocompromised individuals, although knowledge on the reactogenicity and immunogenicity and the impact of prior infection on cellular and humoral immunity in dialysis patients is limited. We now show that dialysis patients mount a robust antibody and T-cell response against the parental SARS-CoV-2 and Omicron BA.4/5 strains targeted by the vaccine, as well as the Omicron BA.1 and BA.2 variants. In line with results from immunocompetent controls, neutralizing activity towards the parental strain was higher compared to Omicron subvariants, whereas T-cell levels towards parental strain and Omicron subvariants were of similarly high magnitude. While IgG-levels, neutralizing antibody activity, and CD8 T-cell levels after vaccination did not differ in patients with and without prior infection, vaccine-induced CD4 T-cell levels were significantly higher in patients with prior infection. Finally, immunogenicity was largely comparable in patients and immunocompetent controls without prior infection. In contrast, among individuals with prior infection, controls had higher neutralizing antibody activity than patients, and dialysis patients had higher CD4 T-cell levels than respective controls.

Our observation that both patients and controls showed lower neutralizing antibody activity towards the Omicron subvariants than towards the parental strain is in line with previous reports on neutralizing antibodies after bivalent vaccination in immunocompetent controls ^15, 17, 32–34^, and with first series of hemodialysis patients ^21, 22^, of which one study also compared antibody responses in patients with and without infection ^21^. Overall, the dominance of neutralizing activity towards the parental strain may result from immune imprinting by previous exposures to the monovalent vaccines and/or to the parental SARS-CoV-2 virus ^35, 36^, which was the dominant strain in our cohort of infected patients. In addition, specific neutralizing activity may further be shaped by infection with SARS-CoV-2 subvariants. While neutralizing activity among non-infected patients and controls did not differ, infected controls who were predominantly infected with BA.2 reached significantly higher neutralizing activity towards parental and Omicron BA.2 than infected patients. Given the similarity in neutralizing antibody response between non-infected patients and controls, it is tempting to speculate whether the higher neutralizing activity among infected controls is mainly driven by the infecting strain. In general, the relative increase in antibody levels and neutralizing activity towards Omicron subvariants in non-infected individuals is more pronounced than in individuals with prior infection, which may suggest that non-infected patients derive greater benefit from bivalent vaccination than infected individuals. Among the infected, the benefit may be stronger in individuals with parental SARS-CoV-2 infection and/or longer distance from infection. When comparing baseline neutralizing activity towards the Omicron subvariants in our cohort of predominantly wildtype-SARS-CoV-2 infected patients with those of a recently published cohort of dialysis patients with a history of Omicron infection ^21^, it seems that our cohort had clearly lower baseline levels and a more pronounced relative increase in neutralizing activity towards Omicron subvariants. Together this may explain why the differences in antibody responses between infected and non-infected patients in our study were less pronounced, and may suggest that the particular benefit of bivalent vaccination for non-infected patients may also extend to infected patients with a history of wild-type infection.

As with antibody levels, percentages of spike-specific CD4 and CD8 T cells after bivalent BA.4/5 vaccination increased in both patients with and without prior infection. However, unlike antibodies, specific T-cell levels against the parental spike protein showed a strong correlation and striking similarity with the vaccine-induced T-cell levels of all tested Omicron VOCs indicating substantial cross-reactivity between the strains. In line with vaccine-induced T-cell responses from immunocompetent controls ^23^, spike-specific T cells were largely polyfunctional, and showed high expression of the immune checkpoint molecule CTLA-4, a marker indicative of recent antigen contact. The higher expression of CTLA-4 in patients without prior infection may indicate some phenotypical evidence of de-novo priming and expansion of a new population of T cells after first contact with BA.4/5 antigen. Consistent with some extent of primary induction ^37, 38^, specific T cells among non-infected patients a restricted cytokine pattern with a lower percentage of multifunctional cells and a relative dominance of dual cytokine-producing cells expressing IL-2 and TNFα, which is different from reactivations, where less multifunctional cells are associated with an increase in cells expressing IFNγ ^37, 38^. As the most striking finding, dialysis patients with prior infection mounted significantly higher levels of spike-specific CD4 T cells than controls with prior infection or non-infected individuals. This finding is compatible with a generally higher disease severity, prolonged disease courses and longer periods of PCR-positivity in immunocompromised patients ^39–42^. As spike-specific T-cell levels in patients with COVID-19 were found to correlate with disease severity ^24^, higher CD4 T-cell levels in dialysis patients may result from more pronounced exposure with viral antigensatthe time of infection. Infected patients were also distinct in that their spike-specific CD4 T-cell levels correlated with those of CD8 T cells, which was not the case among non-infected patients. It therefore seems that induction of T cells by natural infection ^24^ will ensue a more uniform expansion of CD4 and CD8 T cells after subsequent vaccination.

In our study, adverse events of the bivalent BA.4/5 vaccine were collected on a standardized questionnaire, which also inquired which vaccine dose received so far was perceived to be the worst in terms of side effects. It was remarkable that the bivalent vaccine was very well tolerated in both infected and non-infected dialysis patients. As with previous vaccinations, patients either reported no adverse events or primarily local pain at the injection site. This is in line with previous reports of COVID-19 vaccine tolerability and also extends to other vaccine types ^43^. Reactogenicity was significantly different from immunocompetent controls where more individuals reported local and/or systemic reactions that were most frequently pain at the injection site followed by fatigue. The fact that patients rather reported pain at the injection site as compared to systemic adverse events may be due to the fact that polypharmaceutical treatment of patients may have contributed to amelioration of some systemic adverse events. On the other hand, the dialysis procedure itself may be associated with headache and fatigue ^44, 45^; hence some systemic adverse events may not have been perceived as vaccine-related. In any case, in light of our data on the strong immunogenicity, the low rate of adverse events does not seem to correlate with poorer immune responses.

A strength of our study is a detailed analysis of bivalent BA.4-5 vaccine-induced humoral and cellular immunity of dialysis patients, which also assessed the impact of previous infections and a comparison to healthy individuals. Convenience sampling of dialysis patients limits the study to the extent that patients with prior infection were significantly younger than the infection-naïve group. The in part more pronounced immune responses may therefore in part be age-dependent ^46^. However, the control group is matched for age and sex and therefore allows for a direct comparison of the immunogenicity of immunocompetent individuals. We have also not performed any follow-up analyses to assess stability of the bivalent booster response. In light of the more rapid waning of vaccine-induced immunity in dialysis patients ^47^, knowledge on stability will continue to be important to inform future vaccine policies in vulnerable patient groups.

In conclusion, despite insufficient humoral ^6, 7^ and cellular ^8, 9^ immune response compared to healthy controls after the primary COVID-19 vaccine doses, dialysis patients showed a pronounced induction of humoral and cellular immune responses after bivalent booster vaccination. Together with the excellent tolerability, these data are reassuring considering current recommendations towards yearly vaccinations in immunocompromised patients at high risk for severe disease and more rapid loss of specific immunity after vaccination.

## Supporting information

Bronder_supplement

## Supplementary material

Supplementary Figure S1:

Supplementary Table S1: Demographic and clinical characteristics of patients and healthy controls.

## Acknowledgements

The authors thank Christina Baum and the team of the occupational health care center at Saarland University Medical Center, and the team of Department for Kidney Diseases and Hypertension at the SHG clinic in Völklingen for their support in enrolling participants. The authors also thank all participants to this study. Expert technical assistance by Christiane Pallas is acknowledged. Financial support was provided in part by the State chancellery of the Saarland to M.S. Moreover, the work was in part supported by the cluster project ENABLE, the Innovation Center TheraNova, and the LOEWE Priority Program CoroPan funded by the Hessian Ministry for Science and the Arts (HMWK) to M.W.

## Author Contributions

S.B., T.S., J.M. U.S., and M.S. designed the study and the experiments. S.B., R.U., V.K., S.M., C.G., A.A-O., F.H., A.W., M.W. performed experiments. S.B., J.M., U.S., A.A-O., F.H. and M.S. contributed to study design, patient recruitment, and clinical data acquisition. S.B. and M.S. performed statistical analysis. T.S., J.M., U.S. and M.S. supervised all parts of the study. S.B., M.S. wrote the manuscript. All authors approved the final version of the manuscript.

## Data availability

All figures and tables have associated raw data. The data that support the findings of this study are available from the corresponding author upon request.

