## Supplementary material for "Potent induction of humoral and cellular immunity after bivalent BA.4/5 mRNA vaccination in dialysis patients with and without history of SARS-CoV-2 infection": Bronder_supplement

#### Supplementary figure

#### Supplementary figure S1

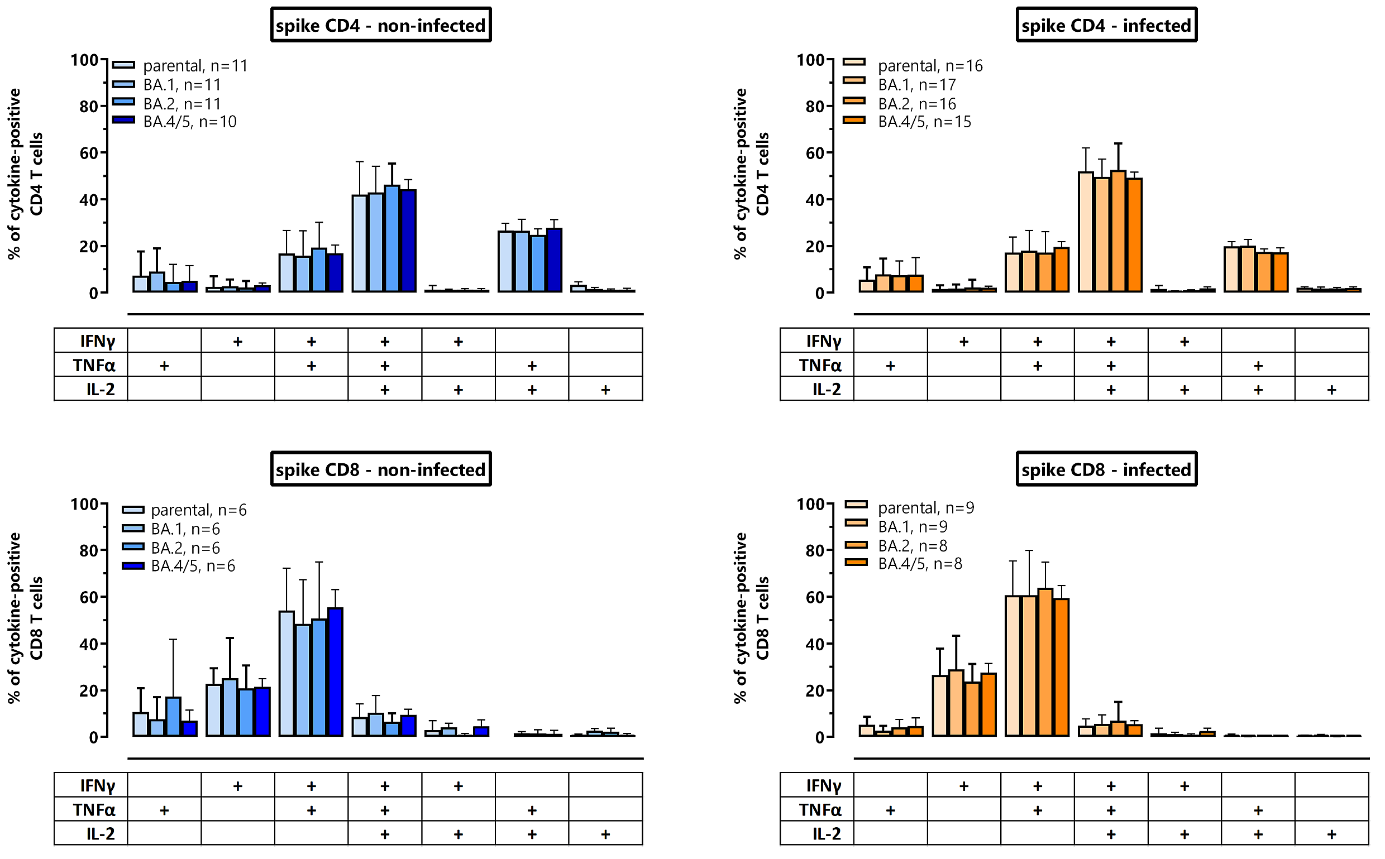

**Supplementary figure S1. Cytokine expression profiles of CD4 and CD8 T cells specific for parental spike and spike of Omicron subvariants.** Comparison of cytokine-expression profile of CD4 and CD8 T cells after respective stimulation with WT-spike as well as the Omicron variants BA.1-, BA.2 and BA.4/5-spike peptides in patients without and with prior infection. Cytokine-expressing T-cells were differentiated into 7 subpopulations according to their expression of IFN-γ, TNF-α and IL-2 (single, double or triple cytokine-expressing cells). Only samples with at least 30 cytokine-expressing CD4 or CD8 T cells were included, respectively. Bars represent means and standard deviations of subpopulations. Differences among subpopulations between the groups were determined using Kruskal-Wallis test with Dunn's multiple comparisons post test. There were no significant differences between the cytokine profiles.

### Supplementary table

#### Supplementary table S1. Demographic and clinical characteristics of patients and healthy controls.

|  |  | non-infected | |  | infected | |  |
| --- | --- | --- | --- | --- | --- | --- | --- |
|  |  | dialysis patients | controls |  | dialysis patients | controls |  |
|  |  | n=14 | n=21 | p-value | n=19 | n=37 | p-value |
| Years of age, mean (SD) | | 71.8 (11.25) | 66.4 (6.6) | 0.082^§^ | 58.7 (15.45) | 55.9 (9.5) | 0.395^§^ |
| Sex, n (%) | | | | |  |  |  |
|  | Female | 7 (50.0%) | 14 (66.7%) | 0.296^†^ | 5 (26.3%) | 21 (56.8%) | 0.051^†^ |
|  | Male | 7 (50.0%) | 7 (33.3%) |  | 14 (73.7%) | 16 (43.2%) |  |
| Vaccine regimen, n (%) | |  |  |  |  |  |  |
|  | homologous mRNA | 13 (92.9%) | 12 (57.1%) |  | 17 (89.5%) | 25 (67.6%) |  |
|  | heterologous | 1 (7.1%) | 9 (42.9%) |  | 2 (10.5%) | 12 (32.4%) |  |
| Infecting strain^$^, n | |  |  |  |  |  |  |
|  | Parental SARS-CoV-2 | n.a. | n.a. |  | 12 | 4 |  |
|  | Delta | n.a. | n.a. |  | 1 | 1 |  |
|  | BA.1 | n.a. | n.a. |  | 2 | 5 |  |
|  | BA.2 | n.a. | n.a. |  | 4 | 21 |  |
|  | BA.4/5 | n.a. | n.a. |  | 0 | 4 |  |
|  | Unknown* | n.a. | n.a. |  | 2 | 3 |  |
| Analysis time  (days after vaccination), median (IQR) | | 16 (2) | 15 (5) | 0.843^§^ | 16 (2) | 14 (2) | 0.183^§^ |
| Differential blood counts  median (IQR) cells/µl) | | n=13 | n=21 |  | n=19 | n=36 |  |
|  | Leukocytes | 6600 (4550) | 6900 (3000) | 0.511^‡^ | 5800 (1900) | 7200 (2050) | 0.015^‡^ |
|  | Granulocytes | 4891 (2536) | 3920 (2465) | 0.807^‡^ | 4437 (1764) | 4267 (1878) | 0.424^‡^ |
|  | Monocytes | 704 (266) | 662 (335) | 0.462^‡^ | 536 (130) | 596 (298) | 0.249^‡^ |
|  | Lymphocytes | 1280 (663) | 2134 (760) | 0.002^‡^ | 1051 (480) | 2297 (700) | <0.0001^‡^ |

^§^unpaired t-test, ^†^Fisher‘s exact test ^‡^Mann-Whitney test; ^ⱡ^information on sex was based on individual self-declaration; ^$^infecting strain based on dominance of SARS-CoV-2 strain at the time of individual infection, 3 individuals had an infection with parental SARS-CoV-2 followed by a second infection with BA.1 (1 patient), BA.2 (1 patient) or BA.4/5 (1 control); *individuals with no known history of infection with a positive NCAP-IgG.
